# Glycemic outcomes of a family-focused intervention for adults with type 2 diabetes: Main, mediated, and subgroup effects from the FAMS 2.0 RCT

**DOI:** 10.1101/2023.09.11.23295374

**Authors:** Lyndsay A. Nelson, Andrew J. Spieker, Robert A. Greevy, McKenzie K. Roddy, Lauren M. LeStourgeon, Erin M. Bergner, Merna El-Rifai, James E. Aikens, Ruth Q. Wolever, Tom A. Elasy, Lindsay S. Mayberry

**Affiliations:** Department of Medicine, Vanderbilt University Medical Center, Nashville, TN, USA; Center for Health Behavior and Health Education, Vanderbilt University Medical Center, Nashville, TN, USA; Department of Biostatistics, Vanderbilt University Medical Center, Nashville, TN, USA; Department of Family Medicine, University of Michigan, Ann Arbor, MI, USA; Department of Physical Medicine and Rehabilitation, Vanderbilt University Medical Center, Nashville, TN, USA; Osher Center for Integrative Health at Vanderbilt, Vanderbilt University Medical Center, Nashville, TN, USA; Department of Biomedical Informatics, Vanderbilt University Medical Center, Nashville, TN, USA

**Keywords:** family, social support, mobile health, randomized controlled trial, type 2 diabetes mellitus, self-care

## Abstract

**Aims:** Family/friends Activation to Motivate Self-care (FAMS) is a self-care support intervention delivered via mobile phones. We evaluated FAMS effects on hemoglobin A1c (HbA1c) and intervention targets among adults with type 2 diabetes in a 15-month RCT.

**Methods:** Persons with diabetes (PWDs) and their support persons (family/friend, optional) were randomized to FAMS or control. FAMS included monthly phone coaching and text messages for PWDs, and text messages for support persons over a 9-month intervention period.

**Results:** PWDs (N=329) were 52% male, 39% from minoritized racial or ethnic groups, with mean HbA1c 8.6±1.7%. FAMS improved HbA1c among PWDs with a non-cohabitating support person (−0.64%; 95% CI [-1.22%, −0.05%]), but overall effects were not significant. FAMS improved intervention targets including self-efficacy, dietary behavior, and family/friend involvement during the intervention period; these improvements mediated post-intervention HbA1c improvements (total indirect effect −0.27%; 95% CI [-0.49%, −0.09%]) and sustained HbA1c improvements at 12 months (total indirect effect −0.19%; 95% CI [−0.40%, −0.01%]).

**Conclusions:** Despite improvements in most intervention targets, HbA1c improved only among PWDs engaging non-cohabitating support persons suggesting future family interventions should emphasize inclusion of these relationships. Future work should also seek to identify intervention targets that mediate improvements in HbA1c.

## 1. Introduction

Despite advancements in type 2 diabetes treatments, approximately half of persons with type 2 diabetes (PWD) do not meet glycemic targets (hemoglobin A1c [HbA1c]) [1-3] with even higher rates among those with minoritized racial and ethnic backgrounds [4 5]. To achieve HbA1c targets, PWDs must regularly engage in diabetes self-care including healthy eating, exercising, stress management, and taking prescribed medication, but adhering to these behaviors is challenging [6 7]. Involving family and friends in PWDs’ self-care efforts may help transform outcomes [8 9]. Family/friends are often present for adults’ daily self-care efforts [10 11] and their support is strongly associated with more regular self-care behaviors and lower HbA1c [12-14]. However, the degree to which engaging family/friends in behavioral interventions for PWDs can improve self-care and HbA1c remains unclear because most interventions have had small and inconsistent effects [15 16].

One reason for inconsistencies could be wide heterogeneity in intervention design and specifically, *how* family/friends are involved [15 17]. Family Systems Theory describes how families, individuals, and health behaviors interact to reinforce and sustain health behavior change [18]. According to the theory, when an individual initiates behavior change, family/friend response will either reinforce or undermine the behavior, affecting the likelihood of maintaining the change [18]. Addressing both the behavior change and the response of family/friends can alter feedback loops and lead to new patterns in the individual’s social system [18]. This theory has been supported in observational studies across chronic diseases, including diabetes [19], but rarely been used to guide interventions.

There is also limited understanding of factors that might explain effects of family-focused interventions on HbA1c [17 20 21]. Although past studies have shown that family-focused interventions lead to improvements in self-efficacy and self-care [15 22], it is not clear whether these factors drive improvements in HbA1c. Furthermore, it remains unclear if improvements in family/friend involvement drive improvements in HbA1c. Finally, average family intervention effects that appear small or null could represent heterogeneity; that is, these interventions may be effective for certain groups and ineffective or detrimental for others [21 23]. For example, gender may play an important role given differences in how men and women want and receive diabetes support [15 24]. Similarly, marginalized groups with less social support [25] may have stronger need or preference for interventions that foster diabetes support from family/friends.

We previously developed and piloted a family-focused, mobile phone-delivered diabetes self-care intervention called FAMS (Family/friends Activation to Motivate Self-care) [26 27]. Informed by Family Systems Theory, FAMS was designed to assist PWDs to initiate self-care goals and learn skills to shape responses (“feedback”) from family/friends. FAMS has several unique attributes. First, FAMS attends to both helpful and harmful responses from family/friends to address inadvertent increases in harmful involvement (e.g., nagging or policing the PWD) due to the intervention [12]. Second, FAMS addresses multiple family/friend relationships for each PWD and gives PWDs the option to invite a support person to receive text messages. Third, FAMS relies only on basic mobile phone technology (i.e., phone calls and text messages) to deliver content, enhancing reach among diverse PWDs [28]. In a 6-month pilot study among a racially and socioeconomically diverse sample, the intervention was acceptable, engagement with each component was high, and FAMS improved family/friend involvement, diabetes self-efficacy, and self-care [27]. Findings from the pilot were used to inform intervention improvements which were usability tested prior to evaluation [29].

The purpose of the current study was to evaluate effects of the improved FAMS intervention on HbA1c among PWDs during a 15-month randomized controlled trial (FAMS 2.0 RCT) [29]. Specifically, we sought to evaluate intervention and sustained effects on HbA1c, as well as examine mediators. We hypothesized that PWDs assigned to FAMS would experience improved HbA1c, diabetes self-efficacy, diabetes self-care behaviors, and family/friend involvement. We further hypothesized that improvements in these intervention targets would mediate FAMS’ effects on HbA1c. Finally, we explored whether intervention effects were different based on PWDs’ gender, racial or ethnic background, socioeconomic status and whether they lived with their enrolled support person.

## 2. Materials and Methods

### 2.1 Study Design

Study procedures were approved by the Vanderbilt University Institutional Review Board and the trial is registered on ClinicalTrials.gov (NCT04347291). Trial design and intervention details have been published [29]. In this 15-month, two-arm RCT, PWDs and their support persons (if enrolled) were randomized to either the 9-month FAMS intervention or enhanced treatment as usual (“control”). PWDs and support persons in both study arms received print materials on managing and providing support for diabetes throughout the 15-month trial; PWDs received text messages advising how to access their study HbA1c results.

### 2.2 Study Participants

Between April 2020 and October 2021, we recruited adults receiving outpatient care for type 2 diabetes at Vanderbilt University Medical Center primary care clinics in Middle Tennessee. Eligible PWDs were between 18 and 75 years of age, able to speak and read English, community dwelling, diagnosed with type 2 diabetes, prescribed at least one daily diabetes medication, and owned a mobile phone. We excluded PWDs with indications of hospice or dialysis services, congestive heart failure, current cancer treatment, pregnancy, dementia, or schizophrenia; or who self-disclosed recent or ongoing emotional, physical, or sexual abuse.

Electronic medical record (EMR) queries identified potentially eligible participants and we prioritized recruitment to patients with most recent HbA1c value ≥8.0% (64 mmol/mol), minoritized racial or ethnic background, and no insurance or public insurance only (proxy for lower socioeconomic status). **Figure 1** details recruitment flow. Of participants contacted and screened for eligibility, 25% (130/519) were ineligible per self-report and 65% (338/519) enrolled; 97% (329/338) of enrolled participants were randomized [29]. Though not required, PWDs were asked to invite a support person, defined as any family member or friend with whom the PWD would feel comfortable talking about diabetes management and health goals.

**Figure 1.**
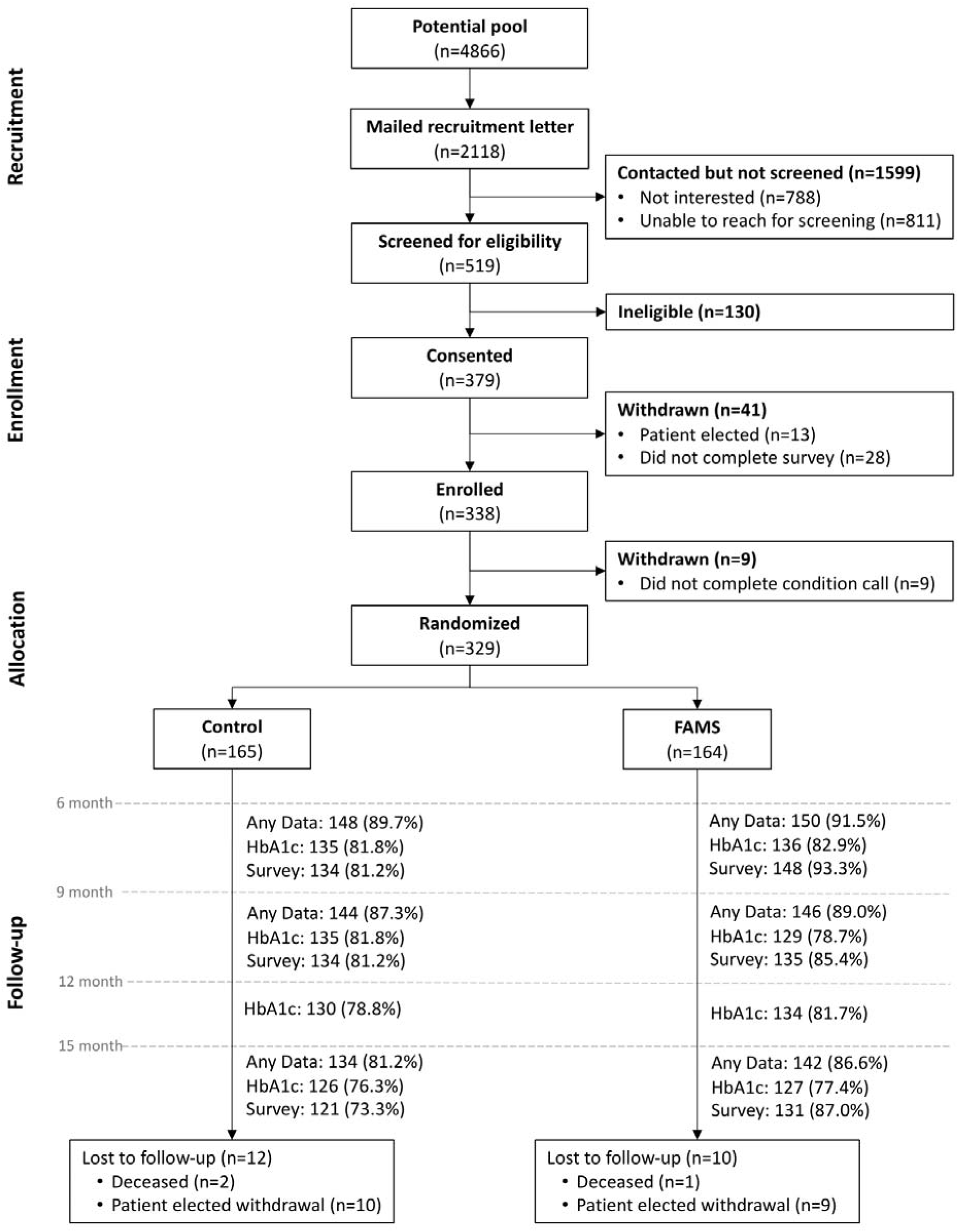
CONSORT flow diagram.

### 2.3 Procedure

After confirming eligibility and providing consent, PWDs (referred to as “participants” henceforth) completed baseline data and were randomized to intervention or control. We administratively withdrew participants who were unreachable for a phone call explaining condition assignment (Figure 1). We used an adaptively stratified randomization process to ensure balance between treatment arms on variables including baseline HbA1c and support person enrollment [29]. Follow-up surveys and HbA1c tests occurred at 6, 9, 12 (HbA1c only), and 15 months. Surveys could be completed via online link, mailed copy, or by phone. HbA1c tests could be completed via venipuncture or point-of-care device at the participants’ clinic or via mail-in HbA1c kits. Participants were compensated up to $200 for completing study measures and all participants received birthday cards and study-branded tote bags. The last follow-up assessment was completed February 2023. All data were stored in REDCap [30 31]. Required data were transferred from REDCap to the digital health platform, PerfectServe, Inc. (Knoxville, TN) to tailor, schedule, and send text messages.

### 2.4 Intervention

FAMS core components [29] include phone coaching and text message support for participants, and text message support for support persons, if enrolled. Monthly phone coaching is designed to improve participants’ ability to identify family/friends’ actions that support or impede self-care goals and learn skills to ask for needed support and manage harmful actions to help meet their goals. Each session begins with behavioral goal setting including a discussion of progress toward the existing goal and then setting a new goal or adjusting the existing goal. Goals can be in the categories of dietary behavior, physical activity, or stress management. Next, participants complete skill-building to improve family/friend involvement relevant to their goal. This includes discussing helpful/harmful involvement and coaches selecting an activity that addresses participants’ needs and experiences. Skill-building activities include observation homework (first session only), activating supports, addressing resistance to involving others, assertive communication, collaborative problem solving, cognitive behavioral coping, and developing an accountability partner. The participant then sets a verbal contract to practice the skill with a specific person and reports on this at the next session. FAMS coaches have master’s level training in helping skills (e.g., social work, counseling, or clinical psychology) and are trained in Family Systems Theory and FAMS coaching protocols.

One-way and interactive text messages are sent to participants and their support persons, if enrolled. Participant texts are dynamically tailored to goals they set during coaching and provide nudges (ideas, tips, encouragement) to engage friends/family in their self-care. Participants also receive texts tailored to self-reported barriers to medication adherence and daily interactive texts asking if they took their medication each day to support adherence. Support person texts seek to increase dialogue about the participants’ goals and facilitate helpful involvement.

### 2.5 Measures

#### 2.5.1 Sociodemographic and clinical characteristics

Participants self-reported demographic characteristics. We created a marker of socioeconomic disadvantage to identify participants meeting any of these criteria: had annual household income <$50,000, public insurance only or uninsured, or ≤a high school degree or GED. We also asked if participants were cohabitating with their support person, if enrolled.

#### 2.5.2 Glycemic outcomes

Our primary outcome was HbA1c collected either from participants’ EMR (venipuncture or point-of-care) or CoreMedica Laboratories mail-in test kit (validated against venipuncture [32]). HbA1c values were included in baseline analyses if taken 42 days before or 21 days after randomization date. HbA1c values were included in follow-up analyses if taken ±42 days of each respective assessment (6, 9, 12, and 15 months from randomization date). When participants had values from both the EMR and a mail-in kit we used EMR values in analyses.

#### 2.5.3 Intervention targets

Intervention targets or hypothesized mediators were diabetes self-efficacy, diabetes self-care behaviors, and family/friend diabetes involvement. Diabetes self-efficacy was assessed with the Perceived Diabetes Self-Management Scale [33]. Each diabetes self-care behavior was assessed using two measures to counter measurement error. Physical activity was assessed with total MET (metabolic equivalent of task) minutes per week calculated using an adapted version of the Rapid Assessment of Physical Activity [34] and with a one-item summative physical activity measure (“Which best describes your current level of physical activity?” with five response options ranging in frequency from “I am very inactive” to “I am active most days”) [35]. Dietary behavior was assessed with the Personal Diabetes Questionnaire scales for problem eating behaviors and use of dietary information for decision making [36].

Medication adherence was assessed with perceived adherence using the reverse-coded Adherence to Refills and Medications Scale for Diabetes [37] and days adherent per week using the Summary of Diabetes Self-Care Activities medications subscale [38]. Finally, we measured helpful family/friend involvement with the Family/friend Involvement in Adults’ Diabetes (FIAD) helpful scale [39] and Important Others Climate Questionnaire [40 41]. We measured harmful involvement with the FIAD harmful scale [39] and an adapted version of the Family Emotional Involvement and Criticism Scale [42 43]. Since both helpful and harmful involvement must be included to estimate effects on HbA1c [39 44], we also calculated a difference score (helpful involvement-harmful involvement) representing overall valence of involvement for mediation models. See Supplementary Table S1 for psychometrics on all self-report measures.

#### 2.5.4 Engagement, study processes, and fidelity

We assessed engagement among intervention participants by calculating response rate to interactive text messages and the average number of coaching sessions completed (1-9, excluding brief wrap-up session 10). Process data characterized coaching sessions, including goal selection across sessions, frequency of goal changes by participant, and skill-building activities used. Coaches entered process data into REDCap within 24 hours of each session. To assess fidelity, coaches used a randomly generated scheme to audio record 40% of sessions (with participant consent) and supervisors reviewed 20% (262 of 1310 completed sessions) against a fidelity rubric. We calculated a fidelity score that ranged from 0-10 with each point representing a step in the coaching session protocol. Supervisors addressed low fidelity scores in biweekly feedback meetings.

### 2.6 Analyses

To estimate FAMS’ main effects and subgroup effects, we used generalized estimating equations (GEE) with a working-independence correlation structure and identity link. Models were adjusted for insulin use at baseline and baseline value of the outcome of interest (via a restricted cubic spline with three knots) for each model. We allowed a two-way interaction between time and condition and a two-way spline interaction between time and baseline value of the outcome. To address missing data, we used multiple imputation via chained equations with M=500 iterations. We had sufficient power to detect a 0.5% difference in HbA1c and standardized effect sizes around 0.3 for intervention targets between conditions at 9 months, as conservative power estimates indicated 80% power for these effects with an effective sample size of 284 [29].

#### 2.6.1 Intervention effects on HbA1c

To assess effects on HbA1c during and post intervention, we used data from imputation models including baseline, and 6- and 9-month assessments. In addition to point estimates, 95% confidence intervals, and *p*-values for 6- and 9-month effects, omnibus tests were performed for joint 6- and 9-month effects. To evaluate sustained effects on HbA1c post-intervention, imputation models included baseline, 12- and 15-month assessments, followed by a GEE model as described above to estimate effects of FAMS on mean HbA1c at 12 and 15 months.

##### 2.6.1.2 Subgroup effects

We stratified the analysis described above by gender (male/non-male), race and ethnicity (non-Hispanic White, non-Hispanic Black, and combined non-Hispanic other races and Hispanic), indication of socioeconomic disadvantage (no/yes), and cohabitation status with support person (non-cohabitating/cohabitating).

#### 2.6.2 Intervention targets and mediation

##### 2.6.2.1 Intervention effects on intervention targets

For each intervention target, we fit models as described above. Continuous measures were analyzed using the GEE model. The summative one-item physical activity measure was ordinal, and therefore analyzed using an ordinal regression model (odds ratios obtained on cumulative logit/proportional odds scale). Each model was run three times, to estimate during (6-months), post (9-months) and sustained (15-months) intervention effects.

##### 2.6.2.2 Mediation of HbA1c effects

Mediation models were estimated using path analysis in Amos version 29. We used Amos’s regression imputation to address missing data, running each imputation separately for each model. We then used imputed data and 2,000 bootstrap samples for bias-corrected estimates and 95% confidence intervals for effects.

Mediation models included hypothesized mediators (i.e., intervention targets) at both 6 and 9 months, and HbA1c as outcome (**Figure 2**). We ran this model separately for HbA1c at 9, 12 and 15 months. For parsimony, we restricted the number of mediators guided by findings evaluating intervention effects on these mediators, opting for continuous and sensitive mediator measure(s) of self-care behaviors unless both measures were significantly improved by the intervention.

**Figure 2.**
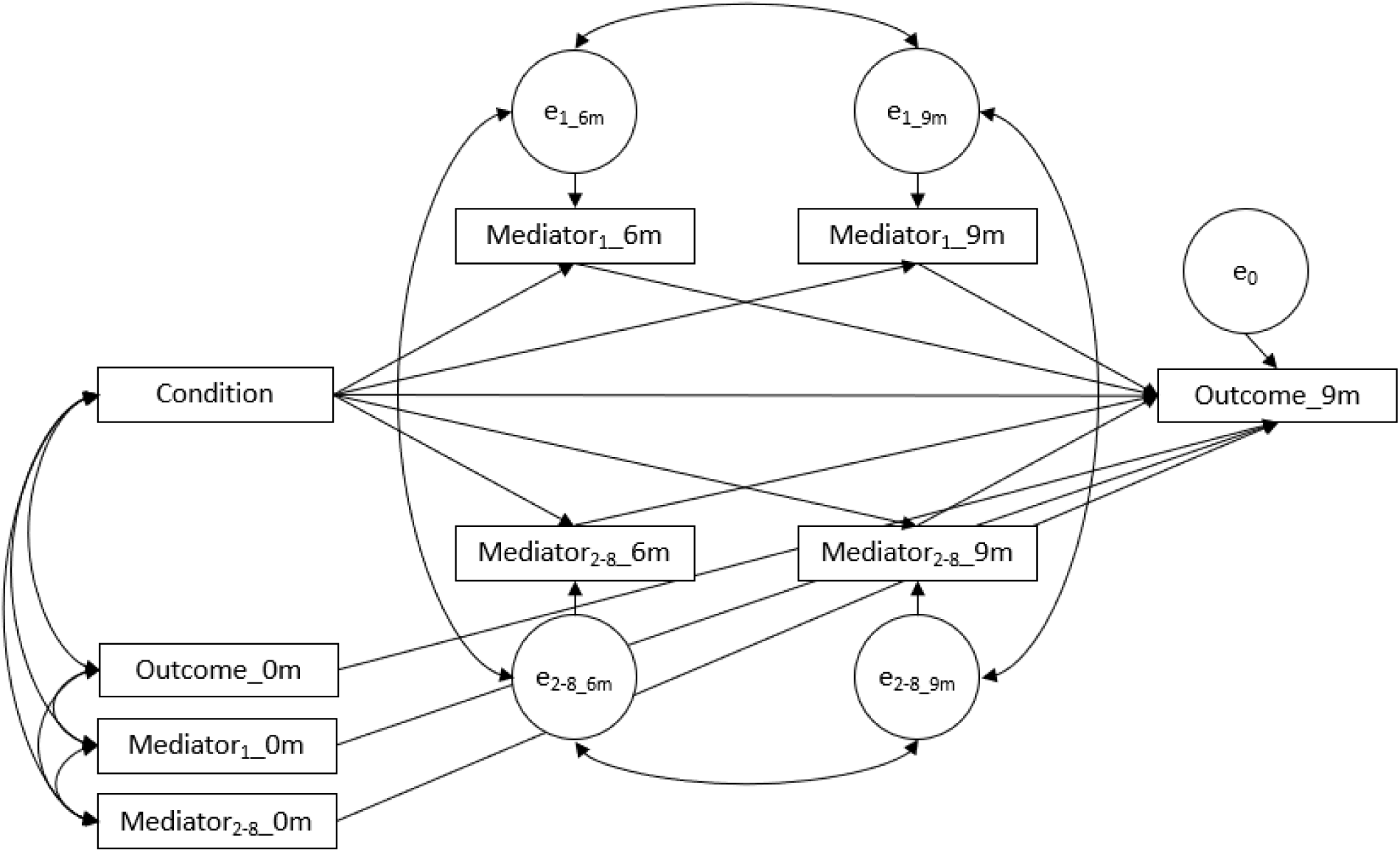
A heuristic depiction of the mediation model predicting hemoglobin A1c (HbA1c) at 9 months. We repeated the model at both 12 and 15 months. For simplicity, the heuristic only shows two mediators; actual models included eight mediators.

Each model was adjusted for baseline values of HbA1c and baseline values of each mediator. Mediators included self-efficacy, self-care (dietary behavior [both aspects: problem eating behaviors and use of information in dietary decision making], weekly MET minutes, and perceived medication adherence), and family/friend involvement (three aspects: involvement difference score [helpful score – harmful score], autonomy support, and perceived criticism). We permitted error variances to covary for 6- and 9-month assessments of each mediator, then across all mediators at the same assessment point (e.g., all mediators at 6 months; Figure 2). We estimated total, direct, and total indirect effects as well as specific indirect effects via each mediator. Specific indirect effects represent the sum of the indirect effect via 6- and 9-month assessments.

## 3. Results

### 3.1 Participant characteristics

N=329 participants (294 with a co-enrolled support person) were randomized. Mean age was 57.0±10.8 years and about half (49%) were female. Over one-third reported a minoritized racial or ethnic background and had annual household income <$50,000. Mean duration of diabetes was 11.5±8.1 years and mean baseline HbA1c was 8.6±1.7% (70±18.6 mmol/mol). Variables of interest were balanced across conditions at baseline (**Table 1**) with all standardized mean differences <0.20 except income and socioeconomic disadvantage, such that intervention participants reported lower income and more socioeconomic disadvantage.

**Table 1.**
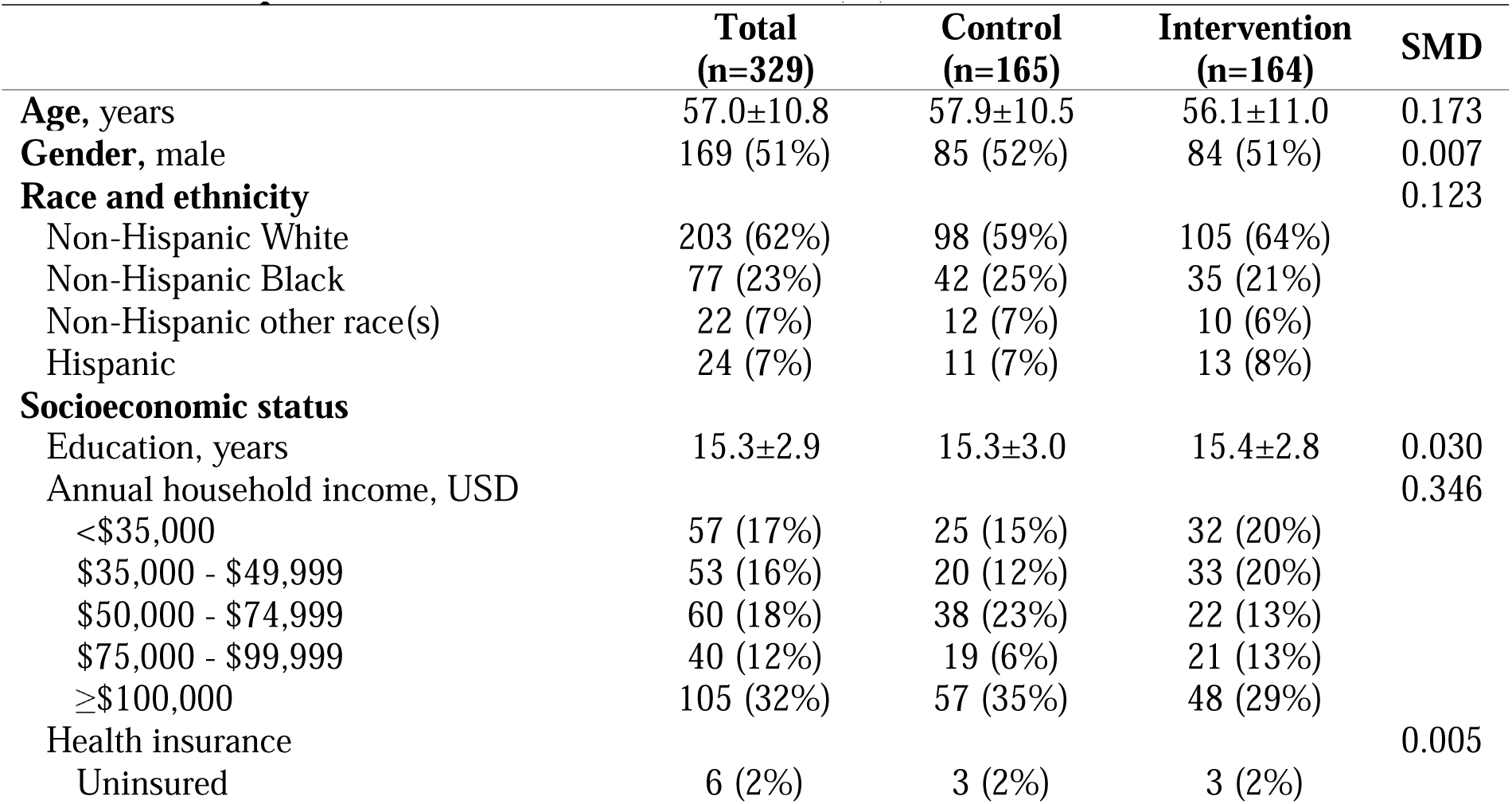
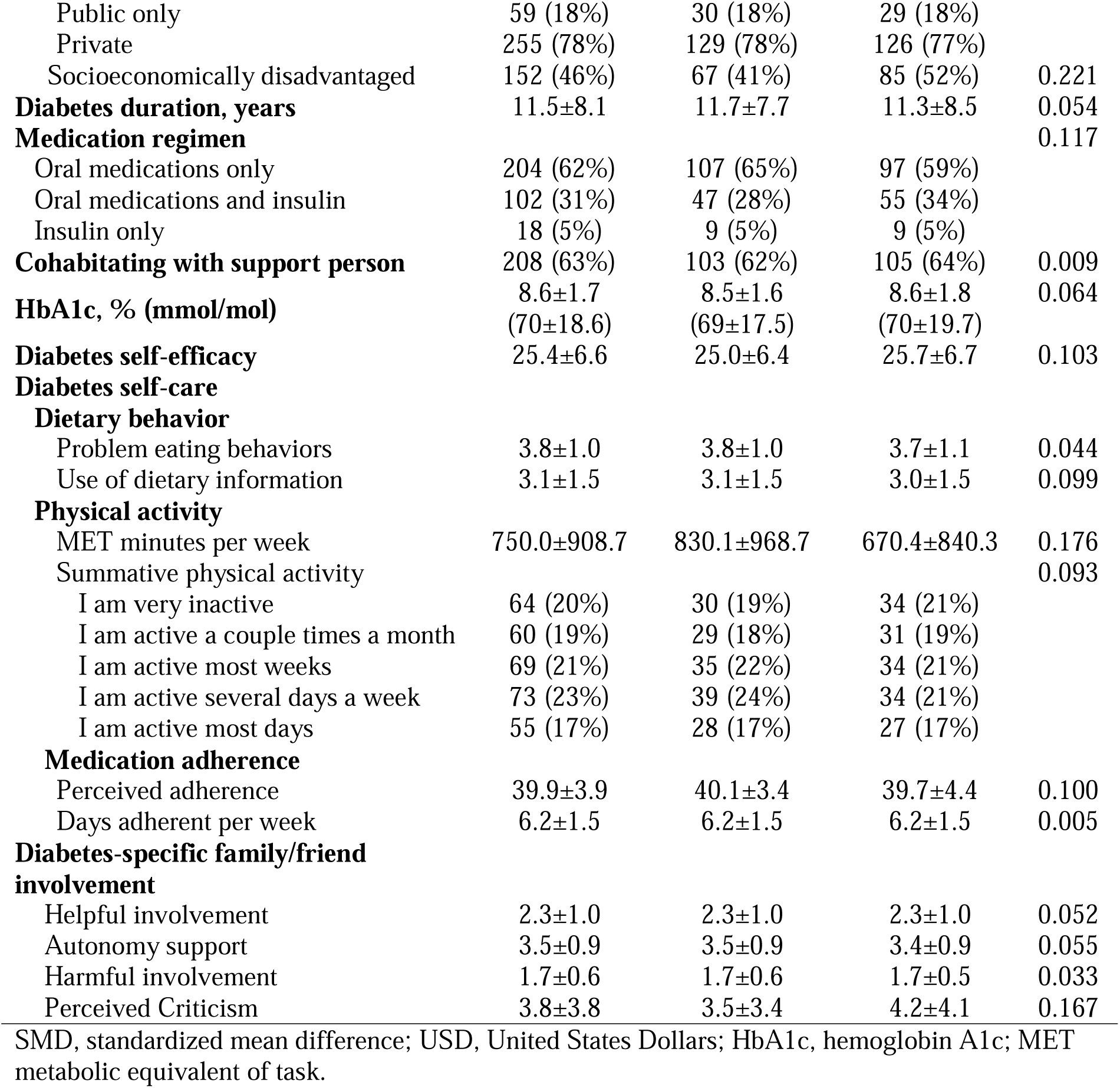
Participant characteristics, Mean ± SD or n(%)

|  | **Total**<br>**(n=329)** | **Control**<br>**(n=165)** | **Intervention**<br>**(n=164)** | **SMD** |
| --- | --- | --- | --- | --- |
| **Age, years** | 57.0±10.8 | 57.9±10.5 | 56.1±11.0 | 0.173 |
| **Gender, male** | 169 (51%) | 85 (52%) | 84 (51%) | 0.007 |
| **Race and ethnicity** |  |  |  | 0.123 |
| Non-Hispanic White | 203 (62%) | 98 (59%) | 105 (64%) |  |
| Non-Hispanic Black | 77 (23%) | 42 (25%) | 35 (21%) |  |
| Non-Hispanic other race(s) | 22 (7%) | 12 (7%) | 10 (6%) |  |
| Hispanic | 24 (7%) | 11 (7%) | 13 (8%) |  |
| **Socioeconomic status** |  |  |  |  |
| Education, years | 15.3±2.9 | 15.3±3.0 | 15.4±2.8 | 0.030 |
| Annual household income, USD |  |  |  | 0.346 |
| <\$35,000 | 57 (17%) | 25 (15%) | 32 (20%) | |
| \$35,000 - \$49,999 | 53 (16%) | 20 (12%) | 33 (20%) | |
| \$50,000 - \$74,999 | 60 (18%) | 38 (23%) | 22 (13%) | |
| \$75,000 - \$99,999 | 40 (12%) | 19 (6%) | 21 (13%) | |
| $\geq \$100,000$ | 105 (32%) | 57 (35%) | 48 (29%) | |
| Health insurance |  |  |  | 0.005 |
| Uninsured | 6 (2%) | 3 (2%) | 3 (2%) |  |
| Public only | 59 (18%) | 30 (18%) | 29 (18%) |  |
| Private | 255 (78%) | 129 (78%) | 126 (77%) |  |
| Socioeconomically disadvantaged | 152 (46%) | 67 (41%) | 85 (52%) | 0.221 |
| **Diabetes duration, years** | 11.5±8.1 | 11.7±7.7 | 11.3±8.5 | 0.054 |
| **Medication regimen** |  |  |  | 0.117 |
| Oral medications only | 204 (62%) | 107 (65%) | 97 (59%) |  |
| Oral medications and insulin | 102 (31%) | 47 (28%) | 55 (34%) |  |
| Insulin only | 18 (5%) | 9 (5%) | 9 (5%) |  |
| **Cohabiting with support person** | 208 (63%) | 103 (62%) | 105 (64%) | 0.009 |
| **HbA1c, % (mmol/mol)** | 8.6±1.7<br>(70±18.6) | 8.5±1.6<br>(69±17.5) | 8.6±1.8<br>(70±19.7) | 0.064 |
| **Diabetes self-efficacy** | 25.4±6.6 | 25.0±6.4 | 25.7±6.7 | 0.103 |
| **Diabetes self-care** |  |  |  |  |
| **Dietary behavior** |  |  |  |  |
| Problem eating behaviors | 3.8±1.0 | 3.8±1.0 | 3.7±1.1 | 0.044 |
| Use of dietary information | 3.1±1.5 | 3.1±1.5 | 3.0±1.5 | 0.099 |
| **Physical activity** |  |  |  |  |
| MET minutes per week | 750.0±908.7 | 830.1±968.7 | 670.4±840.3 | 0.176 |
| Summative physical activity |  |  |  | 0.093 |
| I am very inactive | 64 (20%) | 30 (19%) | 34 (21%) |  |
| I am active a couple times a month | 60 (19%) | 29 (18%) | 31 (19%) |  |
| I am active most weeks | 69 (21%) | 35 (22%) | 34 (21%) |  |
| I am active several days a week | 73 (23%) | 39 (24%) | 34 (21%) |  |
| I am active most days | 55 (17%) | 28 (17%) | 27 (17%) |  |
| **Medication adherence** |  |  |  |  |
| Perceived adherence | 39.9±3.9 | 40.1±3.4 | 39.7±4.4 | 0.100 |
| Days adherent per week | 6.2±1.5 | 6.2±1.5 | 6.2±1.5 | 0.005 |
| **Diabetes-specific family/friend involvement** |  |  |  |  |
| Helpful involvement | 2.3±1.0 | 2.3±1.0 | 2.3±1.0 | 0.052 |
| Autonomy support | 3.5±0.9 | 3.5±0.9 | 3.4±0.9 | 0.055 |
| Harmful involvement | 1.7±0.6 | 1.7±0.6 | 1.7±0.5 | 0.033 |
| Perceived Criticism | 3.8±3.8 | 3.5±3.4 | 4.2±4.1 | 0.167 |

SMD, standardized mean difference; USD, United States Dollars; HbA1c, hemoglobin A1c; MET metabolic equivalent of task.

### 3.2 Engagement, Fidelity, and Study Processes

Over the 9-month intervention period, median response rate to interactive text messages was 82% (IQR: 65%–91%). Average number of coaching sessions completed was 8.2±1.9, with 90% of participants completing 6 or more of 9 sessions. Across all sessions, 32% of participant goals focused on diet, 39% on physical activity, and 29% on stress management. Participants changed goal type across sessions an average of 3.8±1.7 times. Most participants (75%) set each goal type during the intervention. Thirty percent of the coach-selected skill-building activities were collaborative problem solving, 21% activating supports, and 19% developing an accountability partner. Average session duration was 22.6±7.4 minutes (range 8-48) and average fidelity score was 9.4±1.1 (range 2-10); 86% of sessions had fidelity scores score ≥9.

### 3.3 Intervention effects on HbA1c

#### 3.3.1 Intervention effects on HbA1c

At 6 months, the estimated treatment effect on HbA1c trended in the hypothesized direction such that intervention participants had lower mean HbA1c (−0.25%; 95% CI [-0.55%, 0.04%]; *p*=0.090). At 9 months, the estimated effect had diminished (−0.05%; 95% CI [-0.36%, 0.25%]; *p*=0.740). The omnibus test for the 6- and 9-month model was not statistically significant (*p*=0.170). Effects on HbA1c were not sustained at 12 months (−0.02%; 95% CI: [-0.35%, 0.30%]; *p*=0.90) or 15 months (0.02%; 95% CI: [-0.31%, 0.35%]; *p*=0.90).

#### 3.3.2 Subgroup effects for HbA1c

There was evidence of a subgroup effect by cohabitation status such that participants who were not cohabitating with their support person had a significant reduction in HbA1c at 6 months (−0.64%; 95% CI [-1.22%, −0.05%]; *p*=0.033). The magnitude of the estimate at 9 months remained clinically meaningful, though not statistically significant (−0.48%; 95% CI [-1.14%, 0.17%]; *p*=0.15). No other subgroup differences were evident (i.e., gender, race and ethnicity, or socioeconomic disadvantage; see Supplementary Table S2).

### 3.4 Intervention targets and mediation

#### 3.4.1 Effects on intervention targets

**Table 2** reports estimated treatment effects for each intervention target during and post-intervention. We found evidence of intervention effects on diabetes self-efficacy (omnibus *p*=0.006), dietary behavior (problem eating behaviors omnibus *p*=0.009; use of dietary information for decision making at 9 months *p*=0.034) and helpful family/friend involvement (FIAD omnibus *p*<0.001 and autonomy support at 6 months *p*=0.031). We also saw evidence of an intervention effect on physical activity at 6 months using our summative measure that diminished by 9 months; the MET minutes measure indicated potential treatment effects, but large standard errors clouded detection of significance. We did not find treatment effects on harmful family/friend involvement. Effects on intervention targets were not sustained at 15 months (see Supplementary Table S3).

**Table 2.** Effects of FAMS on Intervention Targets.

| Outcome | 6 months |  | 9 months |  | Omnibus |
| --- | --- | --- | --- | --- | --- |
|  | Estimate (95% CI) | *p* | Estimate (95% CI) | *p* | *p* |
| **Diabetes self-efficacy** | 2.15 (0.77, 3.53) | **0.002** | 1.74 (0.37, 3.12) | **0.013** | **0.006** |
| **Dietary behavior** |  |  |  |  |  |
| Problem eating behaviors | -0.27 (-0.45, -0.09) | **0.003** | -0.21 (-0.41, 0.00) | **0.046** | **0.009** |
| Use of dietary information | 0.05 (-0.26, 0.35) | 0.77 | 0.32 (0.02, 0.61) | **0.034** | 0.090 |
| **Physical activity** |  |  |  |  |  |
| Weekly MET minutes | 136 (-48.7, 320) | 0.15 | 174 (-60.5, 410) | 0.15 | 0.22 |
| Summative physical activity<sup>a</sup> | 2.23 (1.43, 3.47) | **<0.001** | 1.08 (0.71, 1.64) | 0.72 | -- |
| **Medication adherence** |  |  |  |  |  |
| Perceived adherence<sup>b</sup> | 0.41 (-0.21, 1.03) | 0.19 | 0.50 (-0.18, 1.17) | 0.15 | 0.28 |
| Days adherent per week | 0.20 (-0.09, 0.50) | 0.18 | 0.14 (-0.24, 0.52) | 0.48 | 0.40 |
| **Helpful family/friend involvement** |  |  |  |  |  |
| Helpful involvement | 0.41 (0.22, 0.60) | **<0.001** | 0.29 (0.09, 0.49) | **0.005** | **<0.001** |
| Autonomy support | 0.19 (0.02, 0.37) | **0.031** | 0.18 (-0.01, 0.37) | 0.060 | 0.074 |
| **Harmful family/friend involvement** |  |  |  |  |  |
| Harmful involvement | 0.02 (-0.10, 0.13) | 0.80 | -0.02 (-0.13, 0.09) | 0.74 | 0.86 |
| Perceived criticism | -0.16 (-0.80, 0.48) | 0.62 | -0.21 (-0.86, 0.44) | 0.52 | 0.79 |

Bolded terms are significant at $p<0.05$ level.
MET, metabolic equivalent of task
<sup>a</sup> One-item measure on an ordinal scale; estimates are adjusted odds ratios. Omnibus test not available for the proportional odds model.
<sup>b</sup> Adherence to Refills and Medications Scale for Diabetes has been reverse-coded such that higher values indicate greater adherence.

#### 3.4.2 Intervention targets mediating HbA1c effects

There was a significant total indirect effect (b=-0.27%, 95% CI [-0.49%, −0.09%], *p*=0.004) on HbA1c at 9 months, suggesting HbA1c reduction was driven by improvements in intervention targets – primarily self-efficacy, dietary behavior and autonomy support (Table 3). The direct effect was significant and positive, indicating the FAMS effect on HbA1c at 9 months was detrimental when mediators were held constant (b=0.32%, *p*=0.017), resulting in a null total effect. At 12 months, the total indirect effect remained significant (b=-0.19%, 95% CI [-0.40%, - 0.01%], *p*=0.043), indicating sustained HbA1c reductions via improvements in intervention targets – primarily dietary behavior (Table 3). However, evidence of sustained indirect effects dissipated by 15 months.

**Table 3.**
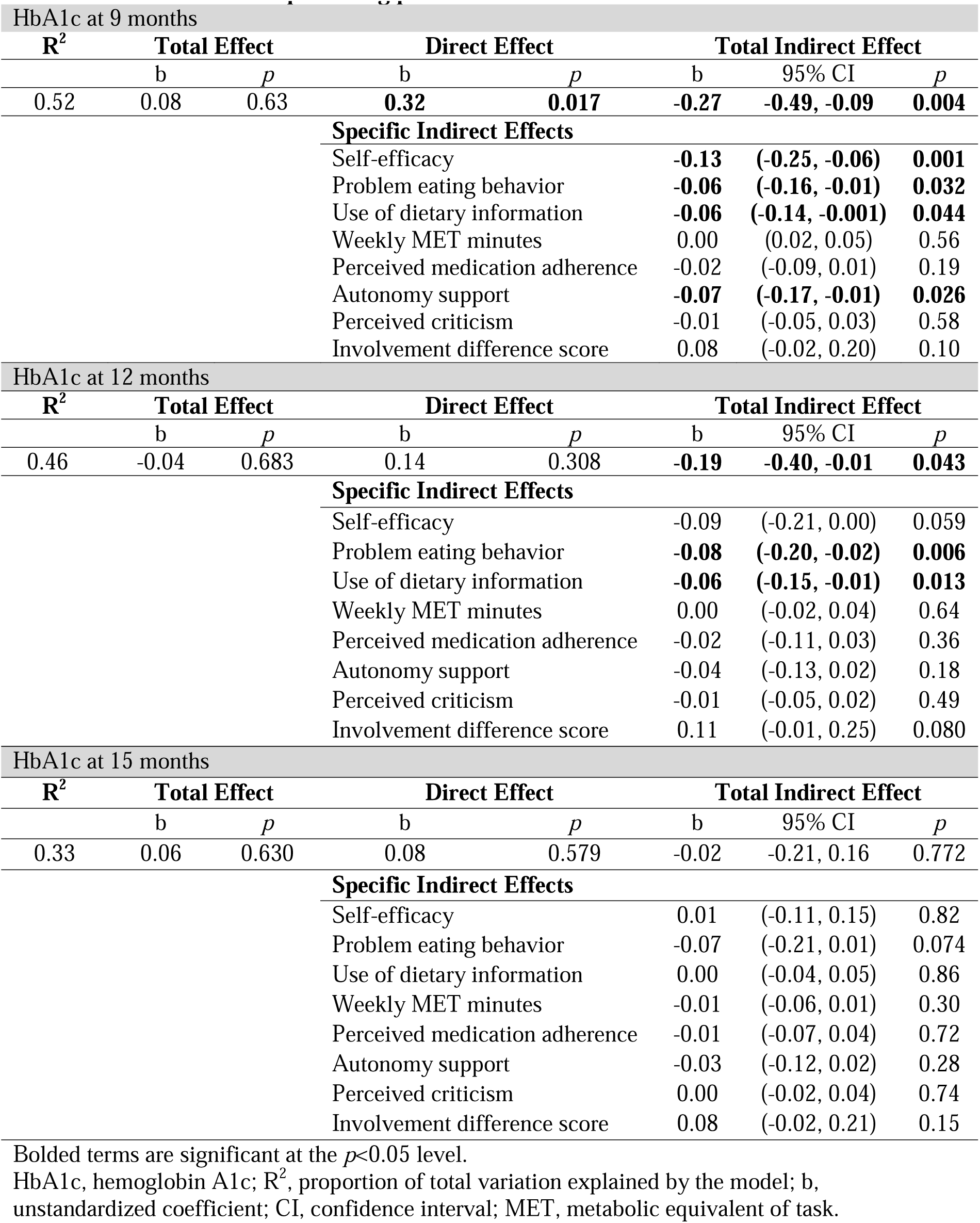
Mediation models predicting post-intervention and sustained HbA1c effects.

## 4. Discussion

We sought to evaluate the FAMS intervention’s effects on glycemic outcomes and address gaps in the literature to advance the science on how and for whom family-focused interventions may improve HbA1c among adults with type 2 diabetes. While there was not sufficient evidence of an overall effect on HbA1c, there was a meaningful HbA1c effect among participants with an enrolled non-cohabitating support person. FAMS improved intervention targets including dietary behaviors, physical activity, self-efficacy, and helpful family/friend involvement. Moreover, HbA1c improvements post-intervention (9 months) and sustained (12 months) were driven by improvements in intervention targets, primarily dietary behavior, self-efficacy, and autonomy support. This is consistent with our conceptual model hypothesizing that improvements in self-efficacy, self-care, and family involvement would drive glycemic improvements, per Family Systems Theory [29]. However, FAMS’ effects on intervention targets and HbA1c all dissipated by the 15-month assessment (6 months post-intervention), emphasizing the ongoing challenge of sustaining behavioral changes.

FAMS’ null mean effects on HbA1c are consistent with the broader literature on family-focused interventions. In a recent trial evaluating another family-focused intervention among adults with type 2 diabetes, Rosland et al. [22] also found improved self-efficacy and self-care behaviors, but not HbA1c. In Zhang et al.’s 2021 meta-analysis of family-based interventions for adults with diabetes, there was an overall effect on HbA1c, but the effect was much larger when the length of follow-up was 6 months or less [45]. This aligns with our study, where HbA1c trended toward significance at 6 months and then dissipated by 9 months.

Our trial advanced knowledge on the effects of family-focused interventions on HbA1c as one of the first studies to examine mediators of improvements in glycemic outcomes. We found evidence of mediated/indirect post-intervention effects and sustained effects 3 months after the intervention ended (at 12 months). However, despite glycemic benefits driven by improved autonomy support, we saw potential (non-significant) harm via other family/friend involvement measures. Harmful family involvement often co-occurs with helpful involvement [12 14] and is more strongly associated with HbA1c observationally [12 44]. Based on existing literature [46], we hypothesized that increasing helpful involvement while *either* reducing harmful involvement or not increasing harmful might result in HbA1c improvements [29]. Our findings suggest reducing harmful involvement may be necessary to improve HbA1c; but reducing harmful involvement is challenging. Zhang et al.’s [45] meta-analysis found significant improvement in helpful involvement but a non-significant improvement in harmful involvement [45]. Finally, FAMS did not improve medication adherence despite tailored and interactive texts supporting adherence. Behavioral interventions may need to improve medication adherence for mean effects on HbA1c given variable glycemic responses to dietary and physical activity changes [47].

We also identified a novel intervention effect in our subgroup analyses; PWDs with a non-cohabitating support person experienced clinically significant HbA1c improvements. This group was heterogenous (living alone, with children only, or with an adult who was not selected to be a support person, possibly due to their own health issues or existing tension/conflict), making it unclear why FAMS effects on HbA1c were strongest in this subgroup. Perhaps out-of-home support extends and strengthens one’s support network or brings less co-occurring harmful involvement. Alternatively, PWDs who did not have a cohabitating support person may have been in greater need of an intervention like FAMS and experienced greater benefit as a result. Regardless, future interventions should be inclusive of out-of-home support persons. Notably, we found no evidence of differential intervention benefit by gender, race or ethnicity, or socioeconomic disadvantage.

Several aspects of the trial enhance confidence in our findings, including meeting recruitment goals, adequate representation of minoritized racial and ethnic groups and persons with socioeconomic disadvantage, high intervention engagement and retention, combined with multiple imputation to reduce bias associated with missing data. However, design choices made toward these strengths come with limitations. First, our EMR-driven recruitment approach successfully represented minoritized and disadvantaged groups to enhance generalizability among important at-risk subgroups, but our sample was drawn from an academic medical center in Middle Tennessee so results may not generalize to other locations. Second, PWDs not interested in family/friend involvement were unlikely to participate (i.e., self-selection bias) – a consistent issue for family interventions. We attempted to reduce this bias by not requiring a support person but doing so introduced heterogeneity of treatment effects. Third, we chose to include HbA1c mail-in kit results and self-reported self-care measures because PWDs who do not regularly come to clinic for HbA1c tests, do not complete dietary recall, and/or do not wear accelerometers are likely different in important ways that can bias results. However, self-report measures are subject to recall and social desirability bias, so we used two measures per behavior and selected scales validated against objective measures when possible. Finally, the intervention was person-centered via PWD-selected goal types, tailoring of skill-building activities, and addressing multiple relationships with family/friends. This approach supports sustainable behavior change [48], but complicates detection of intervention effects because individuals are working toward different behavioral targets.

### 4.1 Conclusions

The FAMS 2.0 RCT demonstrated high intervention engagement and improvements in self-efficacy, dietary behavior, physical activity, and helpful family/friend involvement among a diverse sample of PWDs. FAMS improved HbA1c among PWDs with a non-cohabitating support person, but not overall, suggesting future family interventions should emphasize inclusion of these relationships. Future work should also continue to explore potential moderators of family-focused treatment effects, including aspects of PWDs’ social contexts [43], to identify which PWDs may benefit most from these interventions. FAMS improved intervention targets which mediated improvements in 9-month and 12-month (sustained) HbA1c. However, future interventions that can also reduce harmful family/friend involvement and improve medication adherence may be more successful in improving glycemic outcomes.

## Data Availability

All data produced in the present study are available upon reasonable request to the authors

## Acknowledgements

This research was funded by the National Institutes of Health (NIH), National Institute of Diabetes and Digestive and Kidney Diseases (NIDDK) through R01-DK119282. The content is solely the responsibility of the authors and does not necessarily represent the official views of the NIH. MKR is funded by the Vanderbilt Faculty Research Scholars.

## CRediT authorship contribution statement

**Lyndsay A. Nelson**: Methodology, Writing - Original Draft. **Andrew J. Spieker**: Data Curation, Formal analysis, Visualization, Writing - Review & Editing. **Robert A. Greevy**: Methodology, Writing - Review & Editing, Supervision. **McKenzie K. Roddy:** Investigation, Writing - Review & Editing, Supervision. **Lauren M. LeStourgeon**: Data Curation, Visualization, Project administration, Investigation, Writing - Review & Editing. **Erin M. Bergner**: Writing - Review & Editing, Supervision, Project administration. **Merna El-Rifai**: Project administration, Investigation, Writing - Review & Editing. **James E. Aikens**: Methodology, Writing - Review & Editing. **Ruth Q. Wolever**: Supervision, Methodology, Writing - Review & Editing. **Tom A. Elasy**: Conceptualization, Methodology, Resources, Writing - Review & Editing. **Lindsay S. Mayberry**: Conceptualization, Methodology, Writing - Review & Editing, Formal analysis, Resources, Supervision, Project Administration, Funding Acquisition.

