## Supplementary for "Glycemic outcomes of a family-focused intervention for adults with type 2 diabetes: Main, mediated, and subgroup effects from the FAMS 2.0 RCT"

| Supplementary Table S1: Description and psychometric properties of study measures | | | | |
| --- | --- | --- | --- | --- |
| **Construct** | **Measure** | **Description** | **Validity** ^a^ | **Reliability** ^b^ |
| Diabetes Self-Efficacy | Perceived Diabetes Self-Management Scale (PDSMS) [1] | Eight items assessing confidence in managing diabetes. Responses range from 1=“strongly disagree” to 5=“strongly agree.” Items reverse scored as appropriate and summed to produce a total score ranging from 8 to 40 with higher scores indicating better self-efficacy. | Correlated with self-reported self-care activities and measures of HbA1c [1] | α = 0.87 |
| Physical Activity (MET minutes per week) | Rapid Assessment of Physical Activity (RAPA) [2] | Six items assessing number of days engaged in light, moderate, and vigorous physical activity and amount of time spent in each during a typical week. To increase sensitivity of the measure, we adapted the RAPA to calculate average weekly MET minutes using standard MET minutes for light (3.3), moderate (4) and vigorous activities (8). The equation summed the following for each category of activity: (number of days engaged) x (number of minutes engaged per time) x (MET minutes associated). Total scores range from 0 to 6533 with higher scores indicating more physical activity. | Correlated with caloric expenditure [2] | N/A |
| Physical Activity (Summative  physical activity) | One-item general physical activity [3] | One-item measure asking, “Which best describes your current level of physical activity?” Five response options range in frequency from “I am very inactive” to “I am active most days.” Items treated categorically or continuously depending on analysis. | Correlated with the RAPA per the current study (ρ=0.62, *p*<0.001) | N/A |
| Dietary Behavior | Personal Diabetes Questionnaire (PDQ) dietary behavior scales [4] | Problem Eating Behavior: Three items assessing overeating, unplanned snacks, and poor food choices. Use of Information for Dietary Decision Making: Three items assessing frequency of using information on the number of calories, carbohydrates, and grams of fat in foods to make decisions about what to eat.  For both scales, responses range from 1=“never” to 6=“1 or more times per day;” items reverse scored as appropriate and averaged to create separate subscale scores ranging from 1 to 6. Higher scores indicate more problem eating behaviors and more use of dietary information for decision making. | Correlated with other self-report diet measures [4] and with HbA1c [5] | Problem Eating Behavior: α = 0.81  Use of Information for Dietary Decision Making: α = 0.81 |
| Medication Adherence (Perceived adherence) | Adherence to Refills and Medications Scale for Diabetes (ARMS-D) [6] | Eleven items assessing adherence to diabetes medications. Reverse-scored items as appropriate and summed to produce a total score ranging from 11-44; we reversed the scale so higher scores indicate better adherence. | Validated against other self-report measures and objective refill adherence measures [7]; Independently predicts HbA1c [6] | α = 0.82 |
| Medication Adherence (Days adherent per week) | Summary of Diabetes Self-Care Activities -medications subscale (SDSCA-MS) [8] | Items ask about the number of days in the last week the respondent took the medication, and how many days the respondent took the correct number of doses as prescribed. We asked the medications subscale items separately for oral, injectable, and insulin. Items averaged to produce a total score ranging from 0 to 7; higher scores indicate more days adherent per week. | Correlated with other self-report measures of medication adherence [6]; Independently predicts HbA1c [6] | N/A |
| Family/friend Involvement | Family/friend Involvement in Adults’ Diabetes (FIAD) [9] | Helpful Scale: Nine items assessing helpful family/friend behaviors over the past month.  Harmful Scale: Seven items assessing harmful family/friend behaviors over the past month.  Responses for both subscales range from 1=“never in the past month” to 5=“twice or more each week.” Items are averaged to produce separate scale scores ranging from 1 to 5. Higher helpful scale scores indicate more helpful behaviors from family/friends, and higher harmful scale scores indicate more harmful behaviors from family/friends. | Both subscales correlated with self-care behaviors and HbA1c [9] | Helpful Subscale: α = 0.89  Harmful Subscale: α = 0.60 |
| Family/friend Involvement – Autonomy support | Important Others Climate Questionnaire (IOCQ) [10 11] | Six items assessing family involvement specific to diabetes management. Responses range from 1=“strongly disagree” to 5=“strongly agree.” Items averaged to create a total score ranging from 1 to 5 with higher scores indicating more autonomy supportive communication. | Single factor distinguishable from motivation [10]; convergent validity with other measures of family/friend involvement [9] | α = 0.90 |
| Family/friend Involvement – Perceived Criticism | Family Emotional Involvement and Criticism Scale (FEICS) [12] | Four items assessing perceived criticism specific to diabetes management. Responses range from 0=“almost never” to 4=“almost always.” Items summed to create a total score ranging from 0 to 16 with higher scores indicating more perceived criticism related to diabetes management. | Correlated negatively with communication, problem solving, cohesion, and adaptability [12]; convergent validity with other measures of family/friend involvement [13] | α = 0.85 |

α, alpha; HbA1c, hemoglobin A1c; MET, metabolic equivalent of task; N/A, not/applicable (indicated for single item measures or scales that include items assessing different information for which alpha is not applicable)

^a^ Based on prior studies

^b^ Internal consistency of baseline measure in this study

| Supplementary Table S2. HbA1c effects among subgroups | | | | |
| --- | --- | --- | --- | --- |
|  | **6 months** | | **9 months** | |
| Subgroups | **Est. (95% CI)** | ***p*** | **Est. (95% CI)** | ***p*** |
| **Gender** |  |  |  |  |
| Male | -0.31 (-0.70, 0.08) | 0.12 | -0.14 (-0.54, 0.27) | 0.51 |
| Non-male | -0.20 (-0.63, 0.23) | 0.36 | 0.03 (-0.41, 0.48) | 0.88 |
| **Race and ethnicity** |  |  |  |  |
| Non-Hispanic White | -0.33 (-0.67, 0.01) | 0.060 | -0.14 (-0.50, 0.22) | 0.45 |
| Non-Hispanic Black | -0.07 (-0.69, 0.55) | 0.83 | 0.24 (-0.41, 0.90) | 0.46 |
| Other race(s) or ethnicities | -0.15 (-1.04, 0.74) | 0.74 | -0.10 (-0.95, 0.74) | 0.81 |
| **Socioeconomic Disadvantage** |  |  |  |  |
| Non-disadvantaged | -0.35 (-0.71, 0.02) | 0.064 | -0.16 (-0.56, 0.24) | 0.42 |
| Disadvantaged | -0.21 (-0.70, 0.27) | 0.38 | 0.07 (-0.40, 0.54) | 0.78 |
| **Cohabitation Status** |  |  |  |  |
| Non-cohabitating | -0.64 (-1.22, -0.05) | 0.033 | -0.48 (-1.14, 0.17) | 0.15 |
| Cohabitating | -0.08 (-0.44, 0.27) | 0.64 | 0.24 (-0.11, 0.58) | 0.18 |

| Supplementary Table S3. 15-month effects of FAMS on intervention targets | | |
| --- | --- | --- |
| **Outcome** | **Est. (95% CI)** | ***p*** |
| **Dietary behavior** |  |  |
| Problem eating behaviors | -0.04 (-0.26, 0.18) | 0.74 |
| Use of dietary information | 0.00 (-0.29, 0.29) | 0.98 |
| **Physical activity** |  |  |
| Weekly MET minutes | 65.6 (-151, 282) | 0.55 |
| Summative physical activity ^a^ | 1.11 (0.73, 1.69) | 0.64 |
| **Medication adherence** |  |  |
| Perceived adherence ^b^ | 0.24 (-0.60, 1.08) | 0.57 |
| Days adherent per week | -0.02 (-0.29, 0.26) | 0.90 |
| **Helpful family/friend involvement** |  |  |
| Helpful involvement | 0.04 (-0.16, 0.25) | 0.70 |
| Autonomy support | 0.10 (-0.12, 0.32) | 0.36 |
| **Harmful family/friend involvement** |  |  |
| Harmful involvement | -0.01 (-0.14, 0.12) | 0.86 |
| Perceived criticism | -0.26 (-0.95, 0.42) | 0.45 |
| Bolded terms are significant at *p*<0.05 level.  MET, metabolic equivalent of task  ^a^ One-item measure on an ordinal scale; analyzed using proportional odds model.  ^b^ Adherence to Refills and Medications Scale for Diabetes has been reverse-coded such that higher values indicate greater adherence | | |

R**eferences**
